# A Comparative Study to Find a Suitable Model for an Improved Real-Time Monitoring of The Interventions to Contain COVID-19 Outbreak in The High Incidence States of India

**DOI:** 10.1101/2020.09.14.20190447

**Authors:** G.S Amrutha, Abhibhav Sharma, Anudeepti Sharma

## Abstract

**Background:** On March 11, 2020, The World Health Organization (WHO) declared coronavirus disease (COVID-19) as a global pandemic. There emerged a need for reliable models to estimate the imminent incidence and overall assessment of the outbreak, in order to develop effective interventions and control strategies. One such vital metrics for monitoring the transmission trends over time is the time-dependent effective reproduction number (*R_t_*). *R_t_* is an estimate of secondary cases caused by an infected individual at a time *t* during the outbreak, given that a certain population proportion is already infected. Misestimated *R_t_* is particularly concerning when probing the association between the changes in transmission rate and the changes in the implemented policies. In this paper, we substantiate the implementation of the instantaneous reproduction number (*R_ins_*) method over the conventional method to estimate *R_t_* viz case reproduction number (*R_ins_*), by unmasking the real-time estimation ability of both methodologies using credible datasets.

**Materials & Methods:** We employed the daily incidence dataset of COVID-19 for India and high incidence states to estimate *R_ins_* and *R_case_*. We compared the real-time projection obtained through these methods by corroborating those states that are containing high number of COVID-19 cases and are conducting high and efficient COVID-19 testing. The *R_ins_* and *R_case_* were estimated using R0 and EpiEstim packages respectively in R software 4.0.0.

**Results:** Although, both the *R_ins_* and *R_case_*. for the selected states were higher during the lockdown phases (March 25 - June 1, 2020) and subsequently stabilizes co-equally during the unlock phase (June 1-August 23, 2020), *R_ins_* demonstrated variations in accordance with the interventions while *R_case_*. remained generalized and under- & overestimated. A larger difference in *R_ins_* and *R_case_*. estimates was also observed for states that are conducting high testing.

**Conclusion:** Of the two methods, *R_ins_* elucidated a better real-time progression of the COVID-19 outbreak conceptually and empirically, than that of *R_case_*. However, we also suggest considering the assumptions corroborated in the implementations which may result in misleading conclusions in the real world.

## 1. Introduction

Coronavirus disease (COVID-19) is a severe acute respiratory syndrome caused by a novel Coronavirus (SARS CoV 2). As of March 11, around 53,370 and 4,627 COVID-19 cases and deaths were reported respectively, including 62 cases and 1 death in India.^[1]^ In the light of its rapid spreading in India and an alarming number of daily deaths caused by it, the government of India along with state governments took many strong preventive measures and non-pharmaceutical interventions to flatten the daily incidence and daily death curve. A varying trend of the infection spread have been reported for different locations, which remained ambiguous due to the novel nature of the virus. To unmask this underlying pattern behind the anomalous behavior of the outbreak in response to an intervention, studies to ascertain a suitable epidemiological model are now in the center stage. Accurate modeling of disease transmission is crucial for optimizing control interventions and often-limited healthcare facilities. One of the vital metrics for the assessment of transmission rate over time is the time-dependent effective reproduction number (*R_t_*).^[3]^ *R_t_* is an estimate of secondary cases caused by an infected individual at a time *t* during the outbreak, given that a certain population proportion is already infected. The *R_t_* < 1 indicates the mitigation of the pandemic while a higher *R_t_* i.e. *R_t_*>l, indicates the outbreak progression. A faulty estimate of *R_t_* can systematically under- or overestimate the true transmission rate. Biases are particularly concerning if they occur near the critical threshold viz *R_t_ =* 1. The most prevailing and extensively used methodology for the estimation of the effective reproduction number is proposed by Wallinga and Teunis (2004), often called the case reproduction number (*R_case_*).^[4]^ *R_case_* is basically a probabilistic reconstruction of the transmission network, counting the average secondary cases produced by an individual. However, this method amalgamates the incidence data from time later than *t*, which when applied on a data aggregated on a shorter time step, generates an erroneous *R_t_* estimate producing significantly negative autocorrelation when considered the true incidence.^[5]^ Also, Implementation of *R_case_* sometimes requires sound statistical expertise to produce credible estimates, which is often absent in non-modelers. Previous studies assert that correct modeling is essential in the assessment of policy, population susceptibility and other transmission affecting factors.^[6]^ A fallible *R_case_* estimate of the time-dependent reproduction number could be potentially pernicious. To overcome these shortcomings, Cori et al. proposed the instantaneous reproduction number (*R_ins_*) for real-time estimation of *R_t_* by corroborating incidences that occurred prior to time t, thus indicating a near real-time estimation of disease transmission.^[7]^

Both *R_ins_* and *R_case_* incorporates the generation interval for the estimation of the reproduction number. Precisely, the generation interval is the time interval between the exposure of the infector and the exposure of the infectee. In the case of COVID-19, the accurate time of exposure is extremely difficult to report.^[8]^ The standard proxy for generation time is serial interval viz the time duration between the onset of the symptom in infector and infectee.^[8][9]^ In this paper, we incorporated the serial interval to estimate *R_ins_* and *R_case_*. for India and high incidence states. The study aims to unfold the varying projections of *R_t_* obtained by *R_ins_* and *R_case_* and to probe their credibility by employing the incidence dataset of the states that contain high incidence numbers and has performed robust testing.

## 2. Methods

*R_ins_* and *R_case_* requires the incidence dataset and the serial interval distribution to estimate *R_ins_*. Conceptually different from Cori et al. (2013) and Wallinga et al. (2014), Bettencourt et al. (2008) proposed a methodology to estimate *R_t_* based on the SIR (susceptible-infected-removed) model, that incorporates the growth rate of SIR and estimates the number of infections between the time intervals.^[10]^ Although Bettencourt et al. method has the capacity for a credible real-time *R_t_* estimation, the results could be highly misleading as (1) the model requires a great deal of structural assumptions; (2) SIR based models do not account for the latency period.^[5][11]^ In this work, we focus on *R_ins_* and *R_case_* methods and probe their behavior over the benchmark dataset. The *R_ins_* and *R_case_* estimations were carried out under the standard assumptions: (1) The serial interval of COVID-19 follows a gamma distribution with mean 4 and standard deviation 2. Previous studies investigating the serial interval of COVID-19 validate the same.^[5][12][13]^ (2) The outbreak within the individual state is entirely driven by local transmission. In order to corroborate this assumption, we estimated *R_t_*s (i.e. *R_ins_* and *R_case_*) from the date of the imposition of the first national lockdown viz March 25, 2020. The lockdown as a non-pharmaceutical intervention, resulted in a transmission arrest, ideally confining the outbreak within the state. (3) *R_t_* estimate tends to be misleading in the situation where there is underreporting, however, if the rate of underreporting is constant, then it has no significant impact over the *R_t_* estimate.^[14]^ Sequentially, the rate of underreporting in this study is assumed to be constant.

Here we also analyze the credibility of *R_ins_* and *R_case_* by juxtaposing their respective estimations over two different time periods viz (1) March 25, 2020 to August 23, 2020, and (2) June 1, 2020 to August 23, 2020. The former interval provides an overview of the *R_ins_* and *R_case_* behavior throughout the entire lockdown interventions (Lockdown 1: March 25; Lockdown 2: April 15; Lockdown 3: May 4; Lockdown 4: May 18) and the unlock phases (Unlock 1: June 1; Unlock 2: July 1; Unlock 3: August 23) (detailed description of the initiatives and strategies administered during each phase is given in **Table S2**). The later interval is crucial for a comparative analysis of *R_ins_* and *R_case_* as unlock phases invite abruptions in the infection transmission and we assert that an efficient model should be able to capture these variations. In order to investigate the credibility of the model, the later interval is an ideal window to probe the estimates by the methods.

### 2.1 Dataset

In relevance to the aim of this study, a benchmark dataset is essential to project a true comparison of the reproduction number estimates obtained by the methods. The study corroborates the dataset for the state: (1) If the total incidence reported in a state is ≥ 50,000; (2) If total cases per million in the state are ≥ 1000; (3) If the state has conducted more than 9000 testing per million as on August 23; and, (4) if the percent (%) of positive cases detected among the sample tested (test positive rate TPR) is greater than 5.0 %. This resulted in a sample of eight states namely *Andhra Pradesh, Delhi, Gujarat, Karnataka, Maharashtra, Tamil Nadu, Telangana* and *West Bengal* (**Table 1**). The benchmark dataset of state-wise daily new cases and the daily test counts were obtained from *covid19india.org*. The data of the test sampling efficiency for a state were provided by ICMR (Health departments of State governments).Link to the repository containing all the datasets involved in this study is made available in the additional material.

**Table 1:**
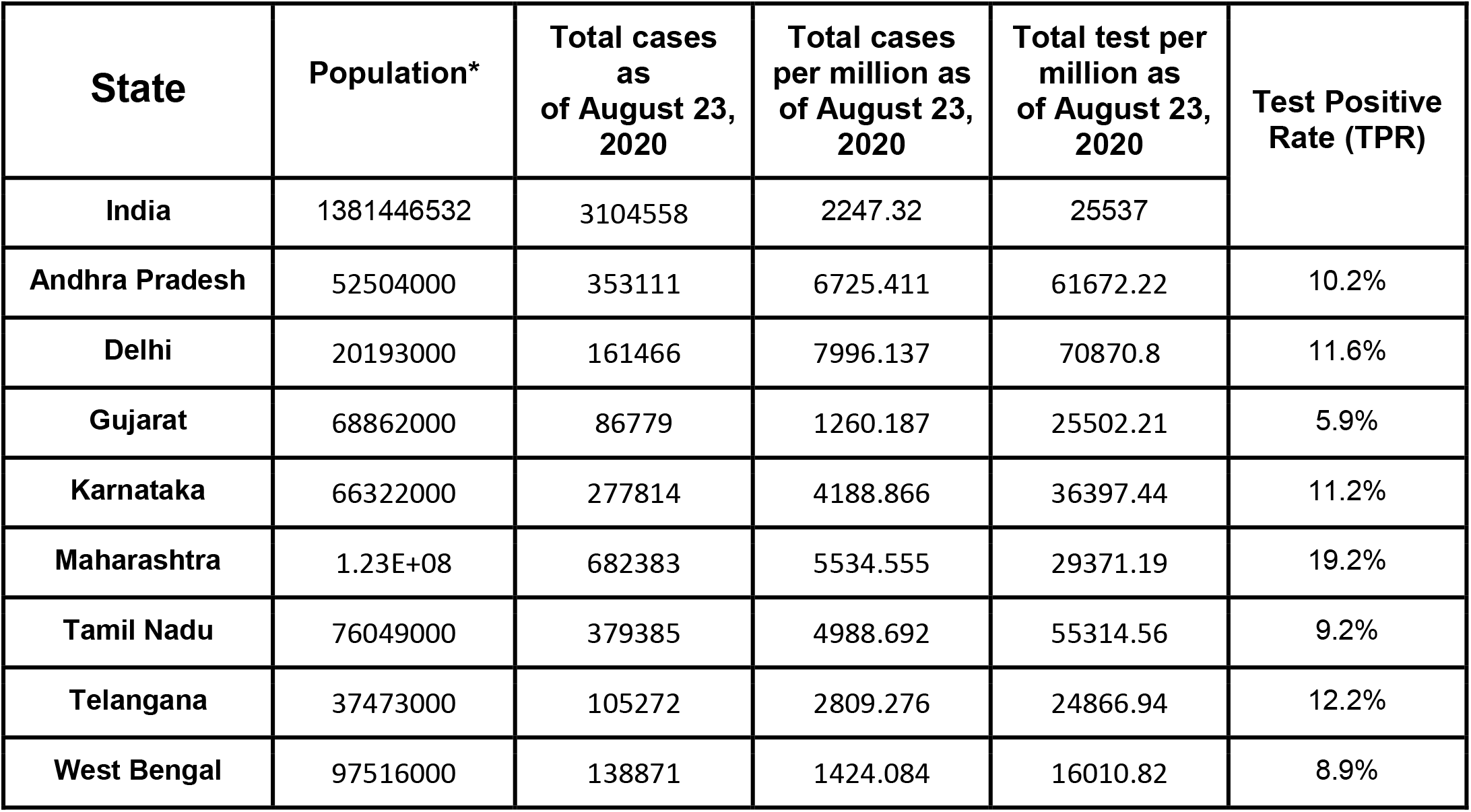
Metrics of India and Indian states containing high incidence of COVID-19. * Projected Total Population as on 1st March - 2020.^[15]^

### 2.2 Case Reproduction Number (*R_case_*)

Wallinga et al. proposed a likelihood-based estimation method to estimate *R_t_*, also called case or cohort reproduction number because it measures the average number of infections that an infected cohort may cause, given that the cohort was infected at time *t*. Precisely, the methods infer “who infected whom” through an observed serial interval distribution while reducing the computational burden by incorporating infector-infectee pairs rather than the complete infection network to measure the likelihood of transmission. Let *t*, denotes the time of the onset of symptoms in the individual *i*. Given a serial interval distribution *w*(*τ*), *τ* being the generation interval, the relative likelihood (*P_ab_*) that individual *a* has been infected by the individual *b* is given as (eq 1):

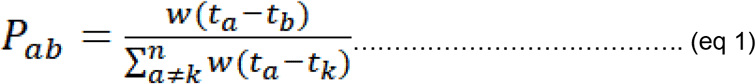

Where *n* is the total incidence reported for each reported case *i*, and *w*(*t*) is the probability of a *t*-day interval i.e. the duration between the onset of symptoms in the infector-infectee pair. The quantity *P_ab_* is a relative likelihood, normalized by the likelihood of the event where *a* is infected by the individual *k*. Sequentially, the Wallinga et al. method obtains the effective reproduction number *R_b_* as (eq 2):

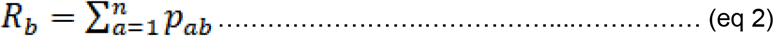

A preeminent overview of the underlying mathematical derivation is described in Wallinga et al. ^[4]^

### 2.3 Instantaneous Reproduction Number

Based on the work of Fraser C. (2007), Cori et al. estimated instantaneous reproduction number *R_ins_* through the ratio of new incidence (*I_t_*) produced at time step *t* to the total infectiousness of infected individuals at time The latter is given as eq 3.

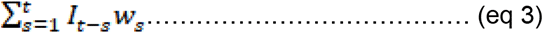

Where *w_s_* is the probability distribution of the infectiousness profile of an individual at *s* units of time from the time of the infection induced in the individual, independent of calendar time *t*. The probability distribution of infectiousness profile depends on biological factors such as immunity or genetics for an individual. Serial interval may be employed as a proxy for *w_s_*. Subsequently, the average incidence at *t* is (eq 4),

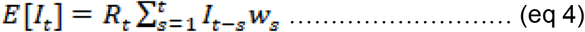

The simulations were performed under the aforementioned assumptions within a confidence interval (CI) of 95%.

## 3. Results and Discussion

The average *R_case_* and *R_ins_* for India during the imposition of Lockdown-1 (March 25, 2020) to the later phase of Unlock-3 (August 23, 2020) was 1.22 (1.19, 1.26; 95% CI) and 1.20 (1.18, 1.21; 95% CI) respectively. Both the metrics were indicating high infection transmission during the early time in the Lockdown-1, consistently declining with two consecutive minor yet significant peaks reported around the end of Lockdown-2 (~ April 30, 2020) and around the beginning of Lockdown-4 (~ May 18, 2020); and eventually continues to stabilize from Unlock-1 and remain steady thereafter (Fig1. A, B). We then compared *R_case_* and *R_ins_* estimates during the unlock phase 1-3 as the state border of the containment zone were sealed but the inter-state movement was unrestricted. The average *R_case_* and *R_ins_* for the post unlock period (i.e. June 1, 2020 to August 23, 2020) was 1.15 (1.13, 1.16; 95% CI) and 1.12 (1.11, 1.12; 95% CI) respectively (Fig1. C, D). The *R_case_* consistently remained higher than average and also elucidated a noisy projection of *R_t_* · *R_case_*. remained high (4.05 (3.77, 4.32; 95% CI) & 3.33 (3.31, 3.36; 95% CI)) early in the time series for both the interval which is very unpragmatic. For entire incidence across India, the two methods almost resembled each other however when estimated over the unlock phase, *R_case_* indicated a significantly higher *R_t_* value of 3.33 (3.31, 3.36; 95% CI) in the beginning of the time interval, which contradicts with its own *R_t_* estimates of 1.14 (1.11, 1.16; 95% CI) when estimated over the entire Lockdown-Unlock phase (i.e. March 25 to August 23) (Fig1. A, C). On the other hand, *R_ins_* estimates for unlock phases remained in line with the estimated *R_ins_* for the entire lockdown-unlock period (Fig 1.B, D).

**Figure_1(A).**
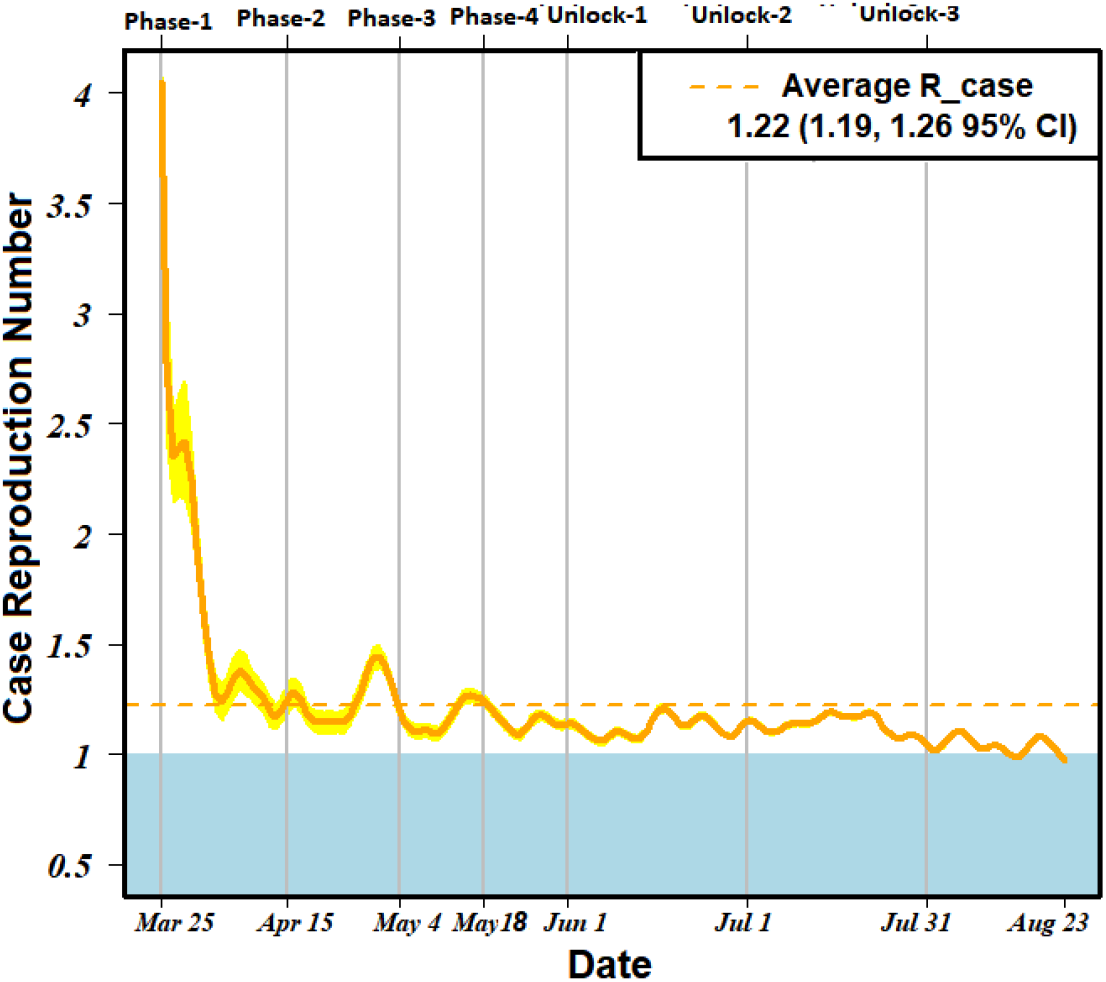
Case reproduction number (R_case) estimate for India from the time of Lockdown-1 till late Unlock-3 (March 25, 2020 to August 23, 2020).

**Figure_1(B).**
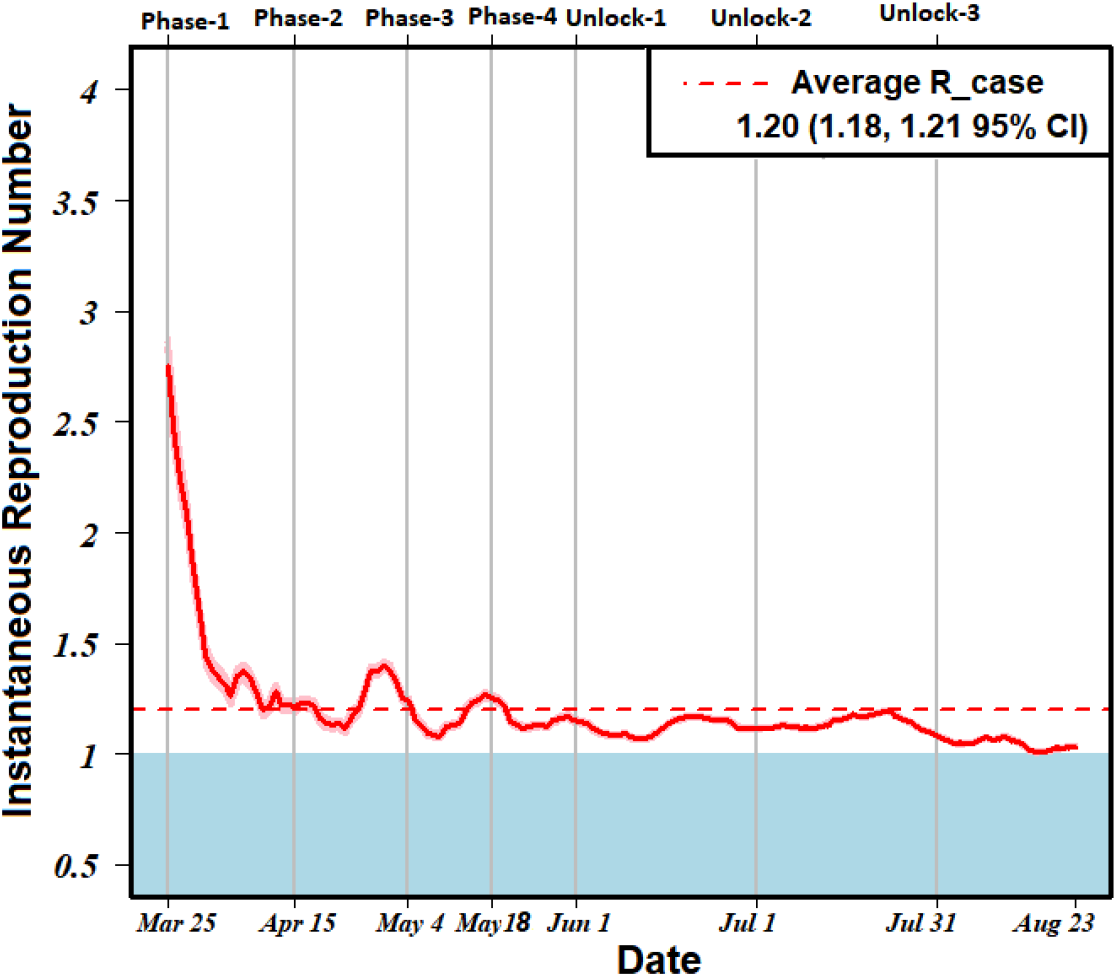
Instantaneous reproduction number (R_ins) estimate for India from the time of Lockdown-1 till late Unlock-3 (March 25, 2020 to August 23, 2020).

**Figure_1(C).**
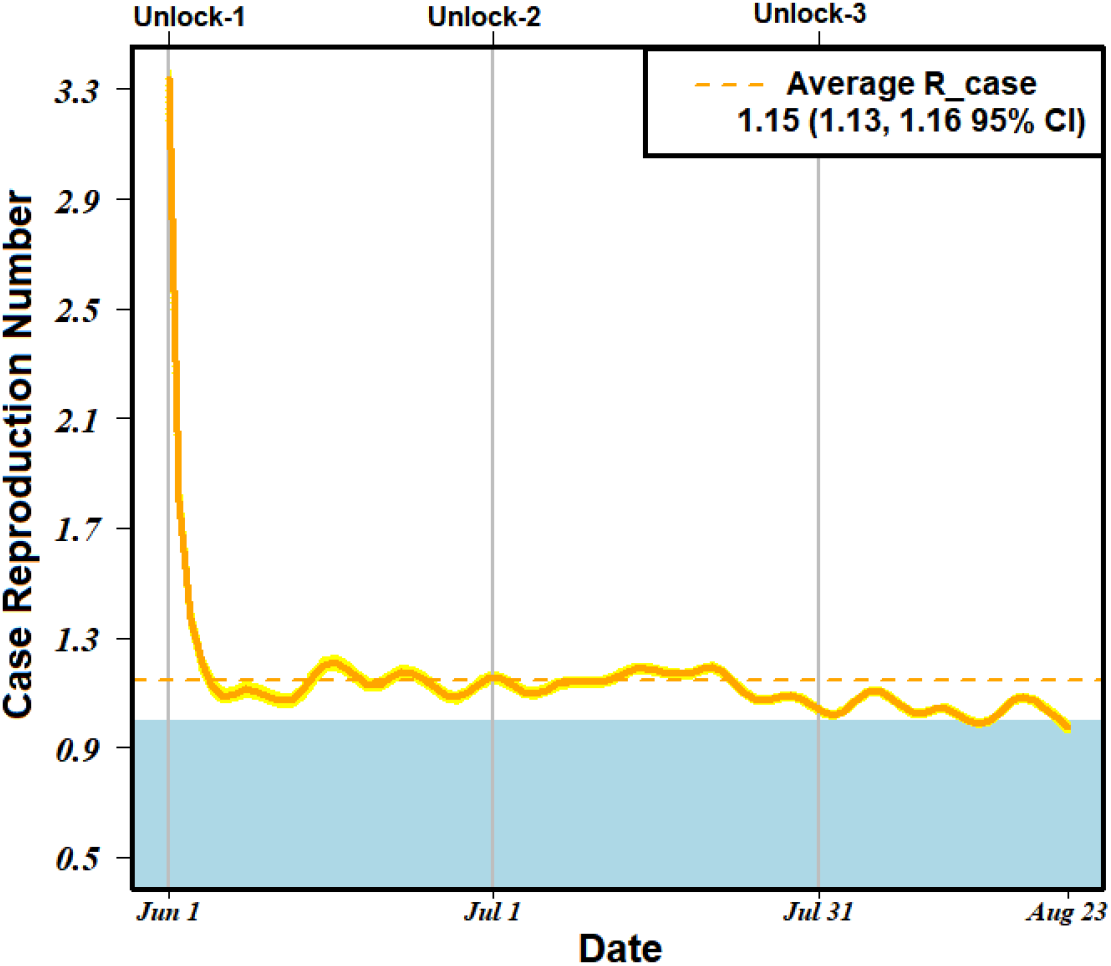
Case reproduction number (R_case) estimate for India from the time of Unlock-1 till late Unlock-3 (June 1, 2020 to August 23, 2020).

**Figure_1(D).**
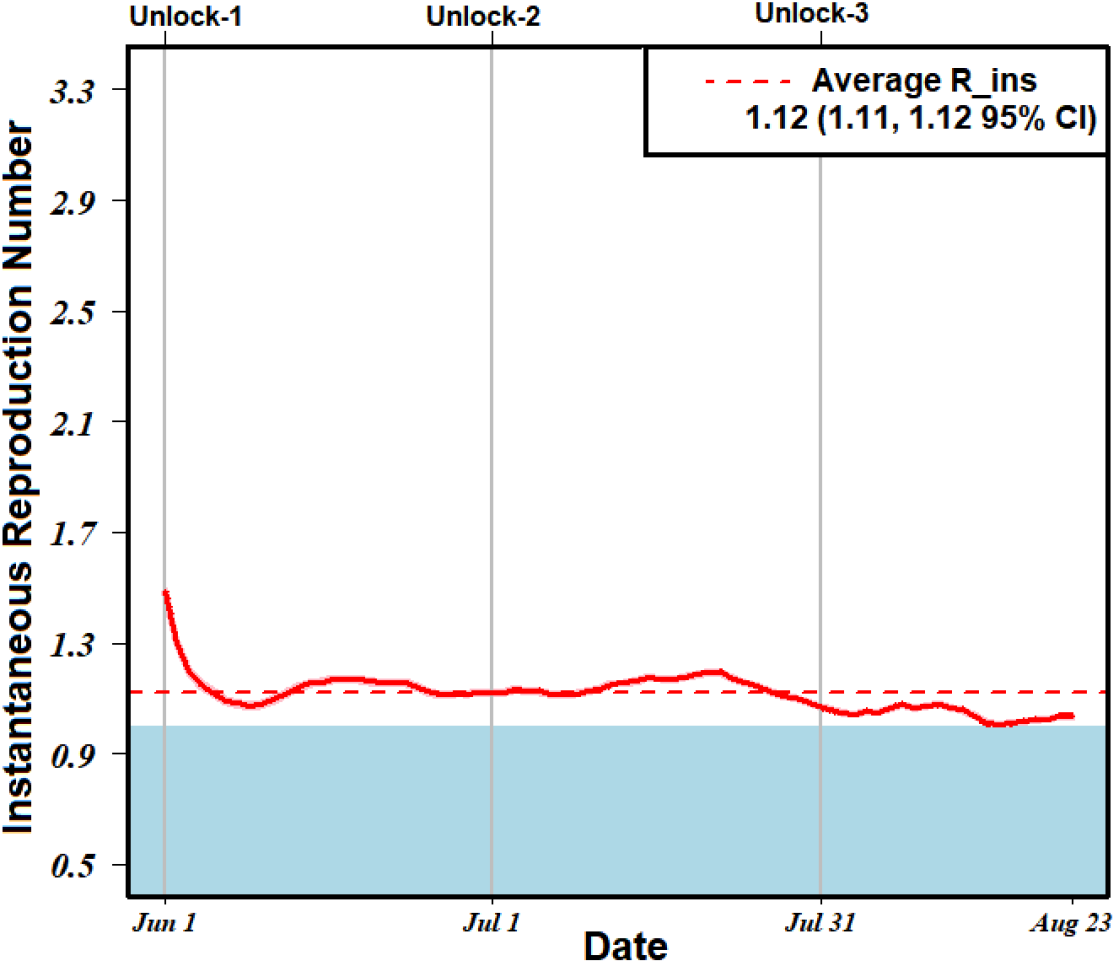
Instantaneous reproduction number (R_ins) estimate for India from the time of Unlock-1 till late Unlock-3 (June 1, 2020 to August 23, 2020).

On the same note, these irregularities in *R_case_* estimates were observed while probing the transmission of COVID-19 in the high incidence states (Fig S1, S2). For each state, the average *R_case_* remained overestimated than average *R_ins_* for both the time intervals viz entire lockdown-unlock duration and the Unlock 1-3. Average estimates of *R_case_* and *R_ins_* for the high incidence states including India is given in Table 2. We found that *R_case_* consistently estimated a high *R_t_* in the initial days of the time series independent of the intervals, which implicates misleading conclusions regarding the infection transmission during the unlock phases (Fig S2). While the real-time estimate of *R_t_* by *R_ins_* was found superior to *R_case_* for all the sample states, this was best illustrated in the cases for Delhi and West Bengal (Fig2). As on August 23, 2020, Delhi contained the highest incidence per million (7996.14) in India and has conducted the highest number of COVID-19 testing per million (70,870.8). The daily new cases in Delhi started to fall significantly from around June 23, 2020, and continued to decline with irregular local minimums (Fig2. A). This consistent fall was best captured by *R_ins_* while *R_case_* remained sensitive for the local peaks and overestimated the incidence during unlock 2-3 (Fig2. B). More importantly, the *R_ins_* detected the variations at earliest while the *R_case_* estimations remained delayed. The underlying mathematical reason behind this anomalistic estimation by *R_case_* is that the *R_case_* amalgamates a weighted sum across transmission events observed after time independent of the trend of transmission before *t*, and this leads to misleading estimations in the latter portion of the time series. While 138,871 total cases of COVID-19 were recorded for West Bengal as of August 23, 2020, the growth of the incidence remained slow during the Lockdown Phase 1-2 viz March 25, 2020 to April 15, 2020, possibly due to underreporting. This anomaly in data was highly underestimated by *R_case_*, indicating a complete end of the transmission (*R_case_* = 0) (Fig2. C), which is very contradictory. On the other hand, *R_ins_* reflected a subtle decline in the estimate but remained in line with the overall true incidence projection by remaining immune to such data inconsistency (Fig2. C). Similar contradictory over- & underestimations by *R_case_* were observed for the cases of other states such as Maharashtra, Gujrat and Telangana (Fig S1, S2).

**Table 2:** Average estimation of case reproduction number (*R_case_*) and instantaneous reproduction number (*R_ins_*) for India and high incidence states for Lockdown-Unlock interval (March 25, 2020 to August 23, 2020) and Unlock 1-2 (June 01, 2020 to August 23, 2020) intervals with 95% CI.

| State | March 25, 2020 to August 23, 2020 |  |  |  | June 01, 2020 to August 23, 2020 |  |  |  |
| --- | --- | --- | --- | --- | --- | --- | --- | --- |
| | Average $R_{case}$ | $R_{case}$ 95% CI | Average $R_{ins}$ | $R_{ins}$ 95% CI | Average $R_{case}$ | $R_{case}$ 95% CI | Average $R_{ins}$ | $R_{ins}$ 95% CI |
| India | 1.22 | (1.19, 1.26) | 1.20 | (1.18, 1.21) | 1.15 | (1.13, 1.16) | 1.12 | (1.11, 1.12) |
| Andra Pradesh | 1.42 | (1.13, 1.74) | 1.28 | (1.20, 1.36) | 1.34 | (1.26, 1.42) | 1.26 | (1.23, 1.28) |
| Delhi | 1.49 | (1.24, 1.73) | 1.19 | (1.14, 1.23) | 1.37 | (1.32, 1.42) | 1.02 | (1.00, 1.03) |
| Gujrat | 1.22 | (1.05, 1.42) | 1.19 | (1.13, 1.26) | 1.09 | (1.02, 1.16) | 1.06 | (1.03, 1.09) |
| Karnataka | 1.27 | (1.02, 1.54) | 1.23 | (1.14, 1.33) | 1.27 | (1.20, 1.34) | 1.20 | (1.18, 1.23) |
| Maharashtra | 1.24 | (1.16, 1.33) | 1.21 | (1.18, 1.24) | 1.13 | (1.11, 1.16) | 1.10 | (1.09, 1.11) |
| Tamil Nadu | 1.37 | (1.20, 1.55) | 1.23 | (1.19, 1.28) | 1.19 | (1.15, 1.22) | 1.10 | (1.08, 1.11) |
| Telangana | 1.28 | (1.03, 1.56) | 1.21 | (1.12, 1.31) | 1.23 | (1.15, 1.31) | 1.17 | (1.14, 1.20) |
| West Bengal | 1.26 | (1.04, 1.51) | 1.23 | (1.14, 1.33) | 1.16 | (1.10, 1.23) | 1.13 | (1.11, 1.16) |

**Figure_2(A).**
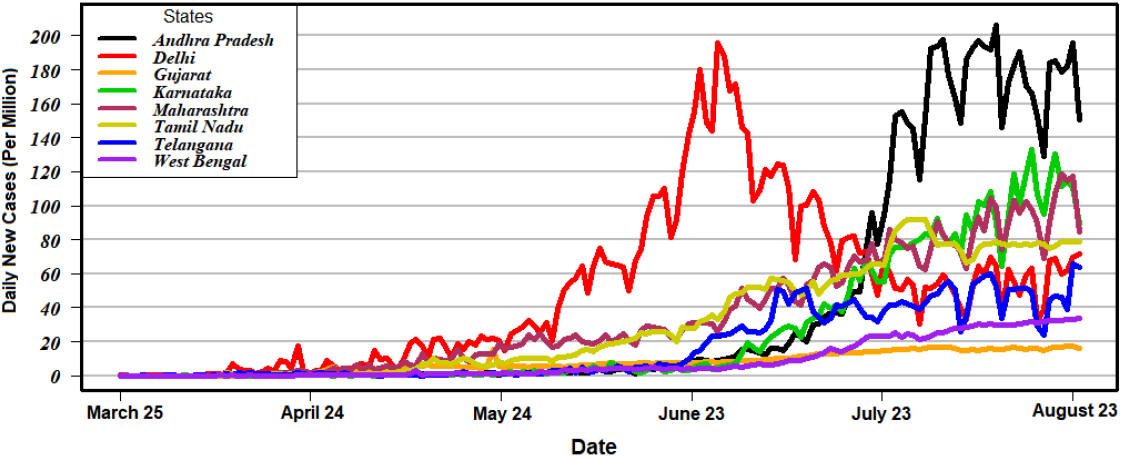
Illustrating daily new cases per million in high incidence states of India from the time of Lockdown-1 till late Unlock-3 (March 25, 2020 to August 23, 2020).

**Figure_2(B).**
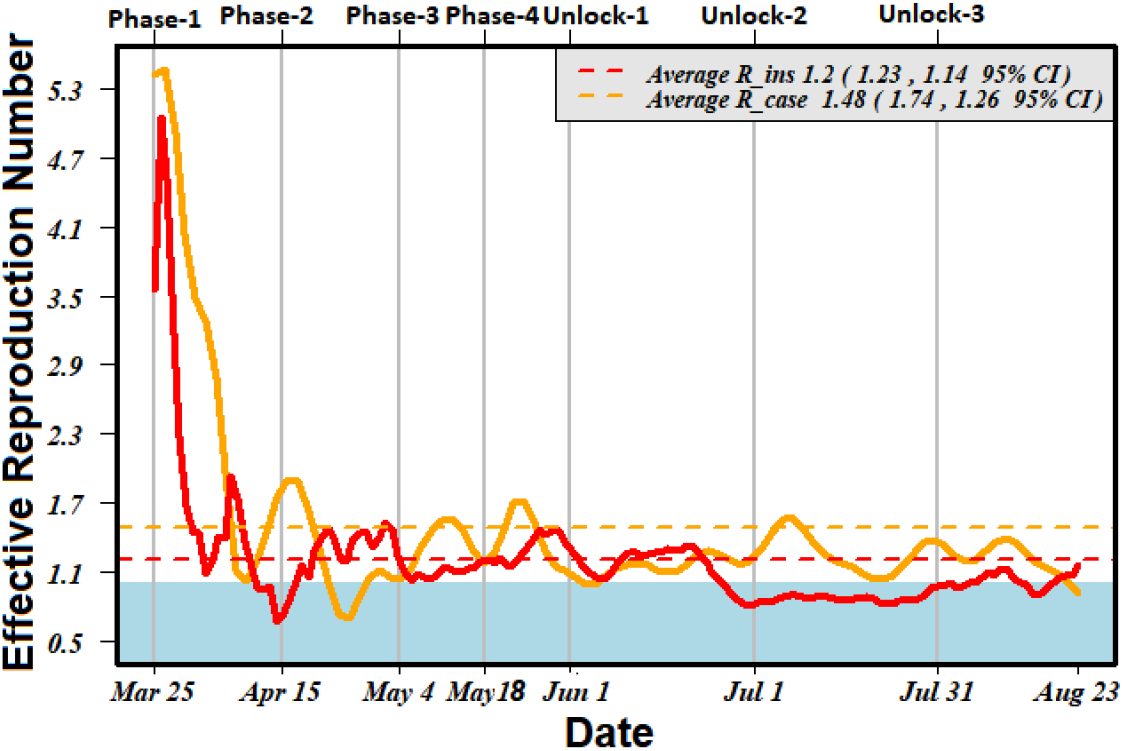
Case reproduction number (R_case) and Instantaneous reproduction number (R_ins) estimation for Delhi from the time of Lockdown-1 till late Unlock-3 (March 25, 2020 to August 23, 2020).

**Figure_2(C).**
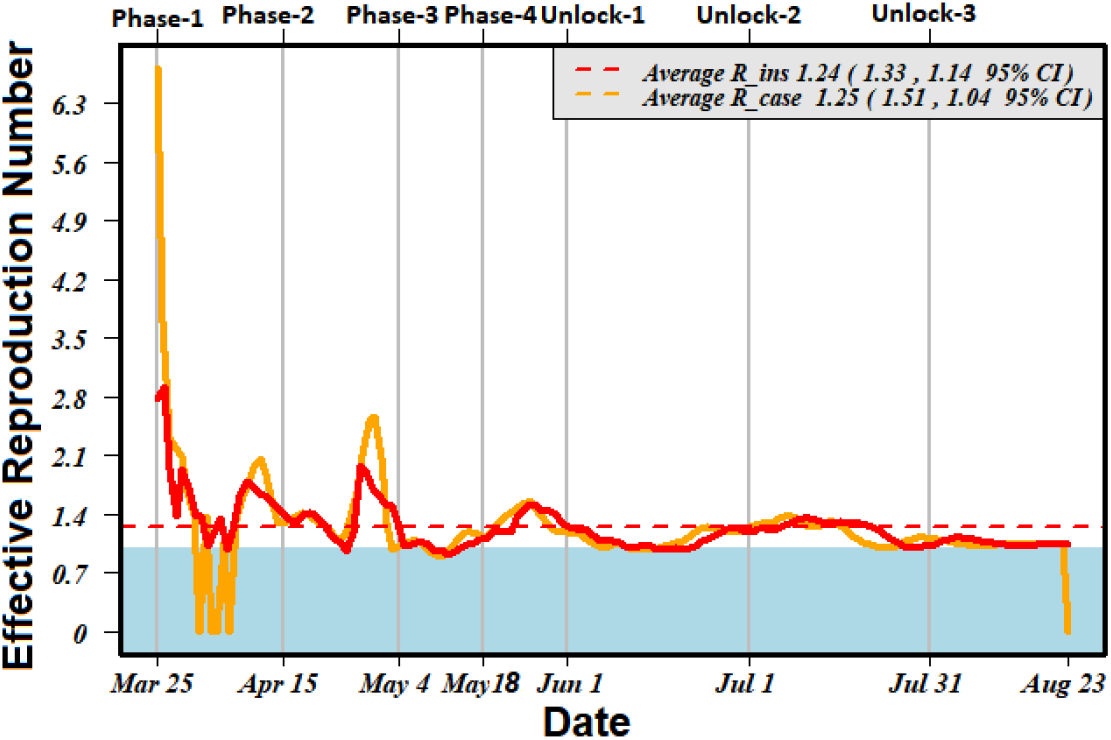
Case reproduction number (R_case) and Instantaneous reproduction number (R_ins) estimation for West Bengal from the time of Lockdown-1 till late Unlock-3 (March 25, 2020 to August 23, 2020).

These deflections of *R_case_* from *R_ins_* estimates for a state were found significantly associated with the amount of testing the state has conducted (Fig3). Higher deflection was observed for the states that have conducted a high number of testing (R square 0.801, p=0.003; CI 95%) (Fig3. B). This association remained intact for the Unlock 1-3 interval as well (Fig3. C). We assert that (1) Because of high testing, the incidence data represents close to real-time incidence occurring in the state, which subsequently facilitates a better comparison of the models. (2) The deflection of *R_case_* from *R_ins_* estimate could also serve as an indicator for assessment of the sufficiency of testing against the infection transmission.

**Figure_3(A).**
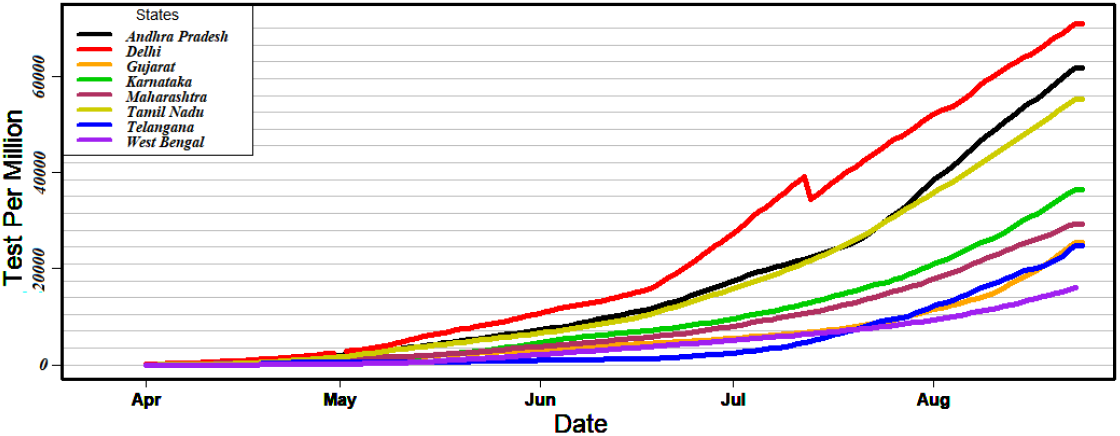
Illustrating daily tests conducted per million in high incidence states of India from the time of Lockdown-1 till late Unlock-3 (March 25, 2020 to August 23, 2020).

**Figure_3(B).**
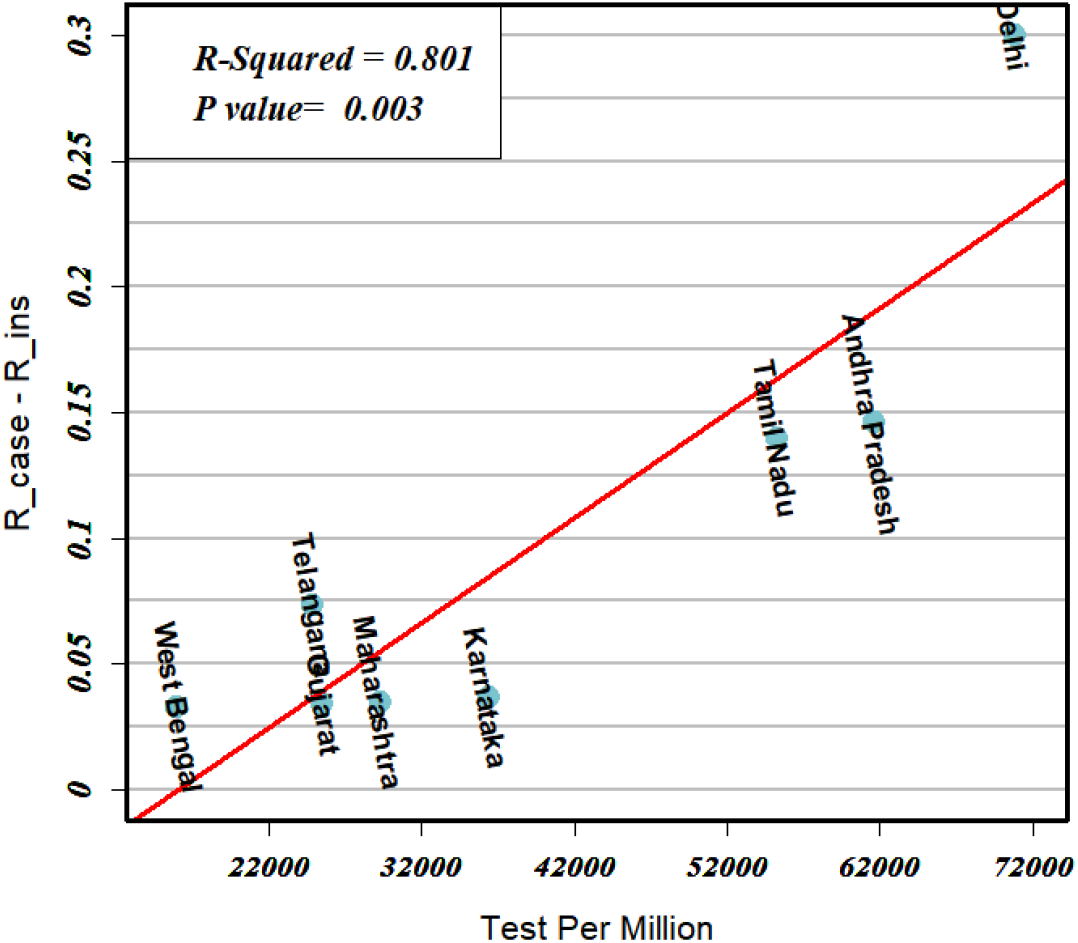
Plots showing the relationship between the difference of and (i.e. R_case - R_ins) measured from the time of Lockdown-1 till late Unlock-3 (March 25, 2020 to August 23, 2020) for the state and the number of testing conducted per million by the state as on August 23, 2020.

**Figure_3(C).**
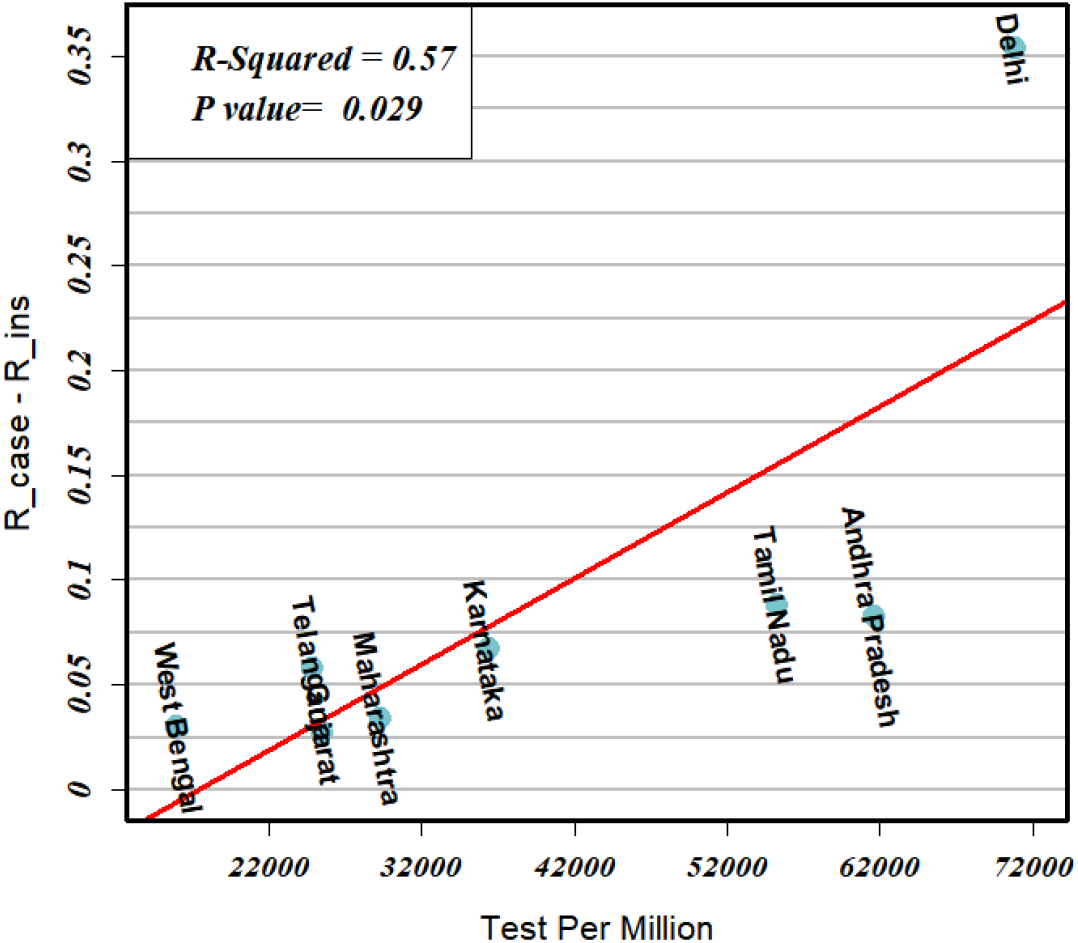
Plots showing the relationship between the difference of and (i.e. R_case - R_ins) measured from the time of Unlock-1 till late Unlock-3 (June 1, 2020 to August 23, 2020) for the state and the number of testing conducted per million by the state as on August 23, 2020.

## 4. Conclusion

Although many studies in the past have employed *R_case_*. for the modelling of effective reproduction number *R_t_* in order to probe COVID-19 outbreak in India, this study retrospectively investigated the contrasting estimates of *R_t_* by *R_case_* and *R_ins_* for the high incidence states of India, during lockdown and unlock phases.^[12][19]^ We found that, (1) while *R_case_*. remained deflected from the true incidence, *R_ins_* captured the variations in line with the real-time trend of incidence recorded in the states during the phases. (2) *R_case_* estimates were lagging from the *R_ins_* values and delayed in the measuring transmission events that occurred days ago. (3) *R_case_* misestimated the transmission rate at beginning of the times while estimations remained credible near the threshold value which is a crucial region to assess the implemented policies.

Since a reliable model is vital for the assessment of the effectiveness of intervention to contain an outbreak, through this work we highly recommend for the implementation of a fine tuned *R_ins_* estimate over *R_case_* to retrospectively monitor the effectiveness of the interventions. However, we also emphasize not to overlook the underlying assumptions made which could invalidate the *R_ins_* estimation and deliver misleading conclusions if not corroborated properly. For instance, in COVID-19 infection, the serial interval distribution might vary from the generation time distribution as the infector can infect the infectee prior to the symptom’s onset, thus indicating a negative value for the serial interval [8]. We hope this work would facilitate an improved assessment modelling for the COIVID-19 outbreak for the optimization and designing of effective interventions to combat the COVID-19 pandemic.

## Data Availability

The source of datasets used for this study are very very well mentioned in the manuscript

https://www.covid19india.org

## 5. Author Contributions

**Amrutha G.S**: Conceptualization, Investigation, Formal Analysis, Writing-Reviewing and Editing. **Abhibhav Sharma:** Conceptualization, Methodology, Data curation, Software, Writing-Original draft preparation. **Anudeepti Sharma:** Formal analysis, Visualization, Writing-Reviewing and Editing.

## Notes

### Competing Interest Statement

The authors have declared no competing interest.

### Clinical Trial

This study is based on the daily new cases of COVID-19 incidence. The data sources are very well mentioned through out the manuscript.

### Funding Statement

No external funding was received

